# Artificial Intelligence–Driven Support and Self-Care Competence as Determinants of Medication Adherence in Diabetes Care: A Cross-sectional Nigerian Study

**DOI:** 10.64898/2026.05.06.26352516

**Authors:** Caleb Onah, Anyalewa Alan Ajonye

## Abstract

Medication adherence among patients with diabetes remains suboptimal in low- and middle-income countries, including Nigeria. Emerging digital health interventions such as AI-powered virtual support may be associated with improved adherence behaviours. This study examined self-care competence and perceived AI-powered virtual support as predictors of medication adherence among patients with diabetes. A cross-sectional survey was conducted among 450 patients recruited through multistage sampling across hospitals in Benue State, Nigeria. Standardised measures of self-care competence scale, perceived AI support scale, and medication adherence scale were analysed using correlation and regression analyses. Results showed that, self-care competence significantly predicted medication adherence (R^2^ = .161), although some components (glucose management, physical activity, healthcare use) showed negative associations. Perceived AI-powered support demonstrated stronger predictive power (R^2^ = .328), with social presence (β = .311, p < .001) and social interactivity (β = .142, p < .01) emerging as key predictors. The combined model explained 36.3% of variance (R^2^ = .363). In conclusion, perceived AI-powered virtual support, particularly socially interactive features, plays a significant role in enhancing medication adherence and may complement traditional self-care strategies. It is recommended that clinicians should therefore adopt a hybrid care model that integrates traditional patient education with AI-assisted interventions. This approach can help bridge gaps caused by high patient loads and limited consultation time, while also enhancing personalised care.

## Background

Diabetes mellitus has emerged as a major public health concern globally, with a rapidly increasing burden in low- and middle-income countries, including Nigeria. Recent estimates indicate that the prevalence of diabetes continues to rise due to urbanisation, lifestyle changes, and demographic transitions, thereby placing significant strain on already fragile healthcare systems (Abdullahi et al., 2025). In Nigeria, the growing incidence of type 2 diabetes mellitus (T2DM) is accompanied by increased morbidity, mortality, and healthcare costs, making effective disease management a national priority. Central to diabetes management is medication adherence, which plays a critical role in achieving glycaemic control and preventing complications such as neuropathy, nephropathy, and cardiovascular diseases (Eze et al., 2022).

Globally, digital health interventions, particularly artificial intelligence-driven systems, are increasingly being integrated into chronic disease management to improve adherence and patient engagement. Evidence from high-income settings suggests that evidence from high-income settings suggests that AI-powered conversational agents… are associated with improved adherence (Abdullahi et al., 2025). However, there is limited empirical evidence on how these technologies function within low-resource contexts, where cultural, infrastructural, and health system differences may influence their effectiveness.

Despite the availability of pharmacological treatments, medication adherence among patients with diabetes in Nigeria remains suboptimal. Empirical evidence suggests that a significant proportion of patients exhibit moderate to poor adherence, resulting in poor glycaemic outcomes and increased risk of complications (Eze et al., 2022). Studies have further shown that adherence is influenced by a complex interplay of socio-demographic, economic, psychological, and healthcare system factors (Onah et al., 2024). For instance, qualitative findings highlight barriers such as financial constraints, poor knowledge of diabetes, lack of family support (Ogwuche et al., 2020), and healthcare system inefficiencies as key determinants of non-adherence (Onwuchuluba et al., 2021; Chinelo & Caleb, 2023). In addition, perceived social support has been identified as a significant predictor of medication adherence among Nigerian patients, suggesting that interpersonal and environmental contexts are crucial in shaping adherence behaviours (Okwuosa et al., 2022).

Self-care competence is another critical factor in diabetes management, encompassing patients’ ability to engage in behaviours such as medication-taking, dietary regulation, physical activity, and blood glucose monitoring. In the Nigerian context, self-care practices have been found to be inconsistent and often inadequate. For example, while medication adherence may be relatively high in some settings, adherence to other self-care components such as diet and glucose monitoring remains poor (Enikuomehin et al., 2021). Evidence from recent Nigerian studies demonstrates that higher levels of self-care competence are significantly associated with better glycaemic control, underscoring the importance of empowering patients with the skills and knowledge required for effective disease management (Abdullahi et al., 2025). However, gaps in patient education, limited access to structured self-management support, and cultural beliefs often hinder the development of adequate self-care competence.

Healthcare delivery systems in Nigeria also face challenges in providing comprehensive diabetes self-management support. Traditionally, the healthcare system has been oriented towards acute care rather than chronic disease management, resulting in insufficient attention to patient education and long-term behavioural support (Iregbu et al., 2022). Healthcare providers often report constraints such as high patient load, limited time, and inadequate resources, which affect their ability to deliver effective self-management interventions. Consequently, many patients are left to manage their condition independently, often without the necessary guidance and support.

In recent years, there has been growing interest in leveraging digital health technologies, including artificial intelligence (AI)-powered virtual support systems, to improve chronic disease management. These technologies offer innovative solutions for enhancing medication adherence through features such as reminders, personalised feedback, and real-time monitoring. Evidence from Nigeria indicates that mobile health interventions can significantly improve medication adherence among patients with diabetes by addressing barriers such as forgetfulness and poor engagement with care (Ihekoronye & Osemene, 2023). AI-powered systems, in particular, have the potential to provide tailored support that adapts to individual patient needs, thereby enhancing self-care competence and adherence behaviours. Thus, the integration of AI-powered virtual support into diabetes care is particularly relevant in resource-constrained settings such as Benue State (Onah & Haruna, 2026), where access to healthcare services may be limited. Virtual support systems can bridge gaps in healthcare delivery by providing continuous, accessible, and cost-effective support to patients outside traditional clinical settings. Furthermore, such technologies can complement existing healthcare services by reinforcing patient education, promoting behavioural change, and facilitating better communication between patients and healthcare providers (Onah & Gwar, 2025).

However, despite the promising potential of AI-driven interventions, there is limited empirical evidence on their effectiveness within the Nigerian setting, particularly in relation to self-care competence and medication adherence. Most existing studies have focused on traditional determinants of adherence, with little attention given to the role of emerging digital technologies. This gap highlights the need for research that examines how perceived AI-powered virtual support interacts with individual factors such as self-care competence to influence medication adherence among patients with diabetes. Given the rising burden of diabetes and the persistent challenges associated with medication adherence in Nigeria, there is a critical need to explore innovative and context-specific strategies for improving patient outcomes. Understanding the combined influence of self-care competence and perceived AI-powered virtual support on medication adherence can provide valuable insights for the development of targeted interventions. Such knowledge is essential for informing healthcare policies and practices aimed at enhancing diabetes management, particularly in underserved regions like Benue State. Beyond addressing an empirical gap, this study extends the Theory of Planned Behaviour by conceptualising AI-powered virtual support—particularly social presence and interactivity—as a form of digital subjective norm. This suggests that AI systems can simulate social influence, shaping adherence behaviours in ways similar to human normative pressures.

Thus, while medication adherence remains a cornerstone of effective diabetes management, it is influenced by multiple factors, including self-care competence and access to supportive interventions. The emergence of AI-powered virtual support systems presents a promising avenue for addressing existing gaps in diabetes care. However, empirical evidence on their role in improving adherence within the Nigerian context remains limited. Therefore, this study seeks to address this gap by examining self-care competence and perceived AI-powered virtual support as predictors of medication adherence among patients with diabetes in Benue State, Nigeria.

### Research Questions

The following research questions for this study to:

i. To what extent does self-care competence predict medication adherence among patients with diabetes in Benue State?
ii. To what extent does perceive AI-powered virtual support predict medication adherence?
iii. To what extent do self-care competence and perceived AI support jointly predict medication adherence?

### Hypothesis of the Study

The following hypothesis were formulated to guide this study;

i. Self-care competence will significantly predict treatment adherence among diabetic patients in Benue State Hospitals.
ii. Perceived AI support will significantly predict treatment adherence among diabetic patients in Benue State Hospitals.
iii. Self-care competence and perceived AI support will jointly predict treatment adherence among diabetic patients in Benue State Hospitals.

**Diagram 1:**
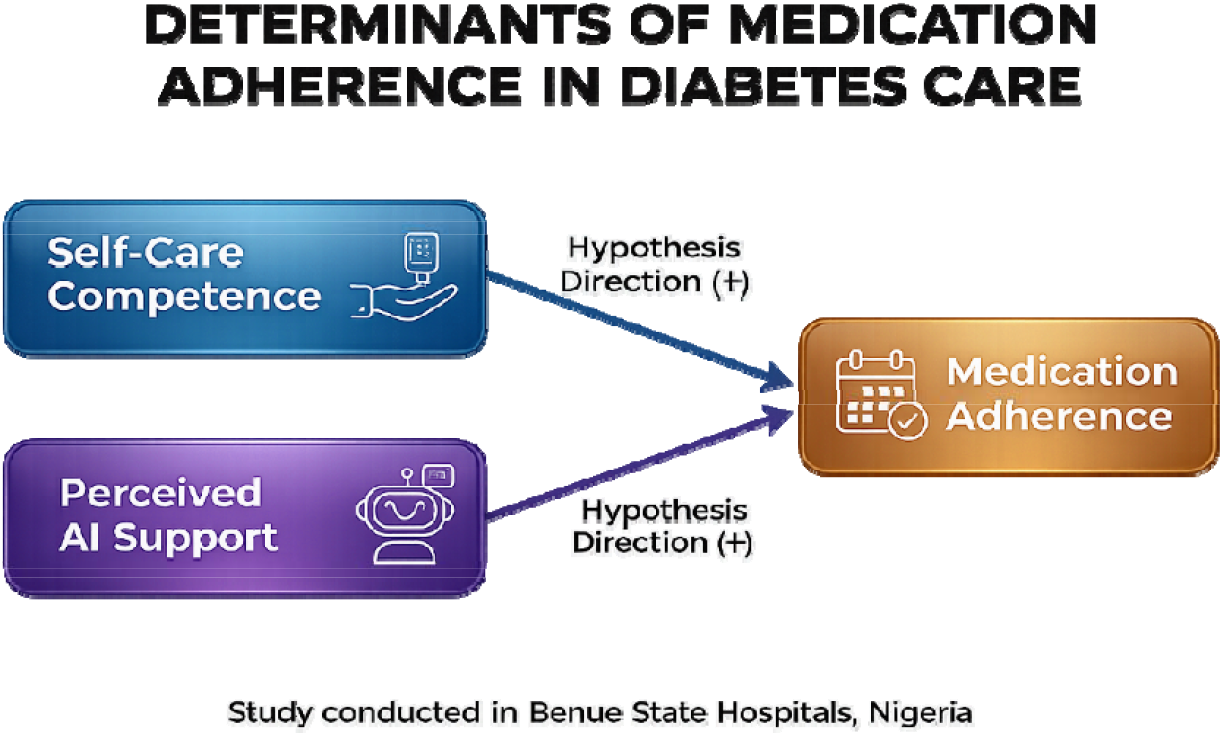
Hypothesised Path Analysis: Self-Care Competence, Perceived AI Support, and Medication Adherence.

### Theoretical Framework

This study is anchored on the Theory of Planned Behaviour (TPB) proposed by Ajzen (1991). The TPB posits that behaviour is primarily predicted by behavioural intention, which is influenced by three constructs: attitude toward the behaviour, subjective norms, and perceived behavioural control (Ajzen, 1991). In the context of this study, self-care competence aligns strongly with perceived behavioural control. Patients who possess higher competence in glucose monitoring, diet, and physical activity perceive themselves as more capable of managing diabetes. This perceived control can enhance or, as your findings suggest, sometimes complicate adherence, particularly when patients substitute lifestyle management for medication (Abdullahi et al., 2025).

**Diagram 2:**
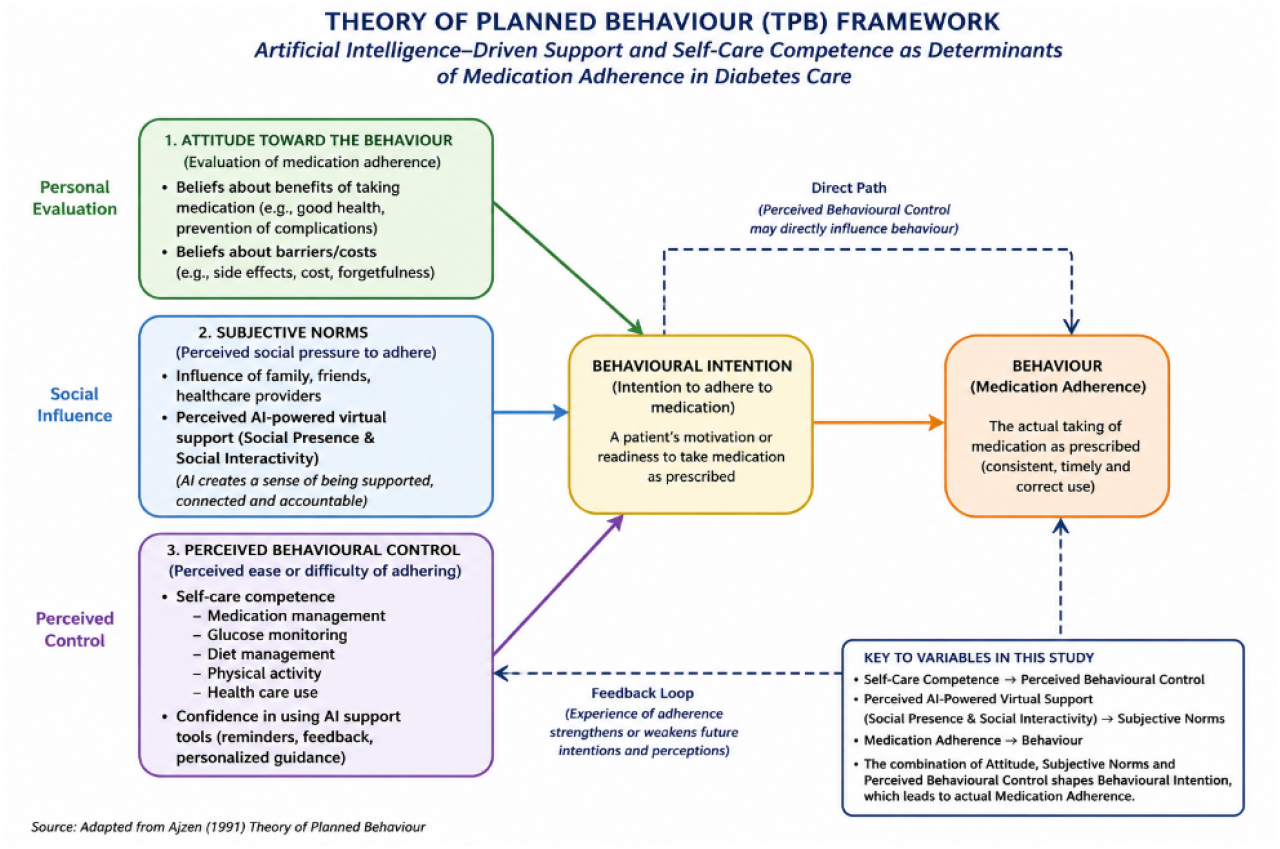
Pathway of the Theory of Planned Behaviour (TPB) proposed by Ajzen (1991)

Perceived AI-powered virtual support fits within both subjective norms and perceived behavioural control. Socially interactive AI features—such as social presence and interactivity—simulate human support, thereby creating a sense of accountability and encouragement. This reflects subjective norms, where patients feel “supported” or “observed,” strengthening adherence intentions (Ihekoronye & Osemene, 2023). Simultaneously, AI systems provide reminders, feedback, and personalised guidance, enhancing patients’ confidence in managing their treatment, thus reinforcing perceived behavioural control (Onah et al., 2024).

Medication adherence, the outcome variable, represents the actual behaviour resulting from these cognitive and social processes. The stronger predictive power of AI support observed in your study suggests that technologically mediated social influence may be particularly salient in the Nigerian context, where healthcare system constraints limit continuous human interaction. TPB effectively integrates both internal (self-care competence) and external (AI-driven support) determinants of adherence. It explains not only why patients intend to adhere but also how digital health innovations modify behavioural pathways. Thus, the theory provides a robust framework for understanding the combined and interactive effects of psychological competence and AI-enabled support in diabetes care within Nigeria.

This study further extends TPB by proposing that AI-driven social presence represents a form of digitally mediated subjective norm, indicating that normative influence can emerge not only from human agents but also from interactive technological systems.

## Methods

### Participants and Recruitment

This study employed a cross-sectional survey design involving 450 patients with diabetes (response rate = 90%) recruited through a multistage sampling procedure across 20 hospitals in Benue State, Nigeria. Participants includes both in-patients and out-patients who meet the following inclusion criteria; a clinical diagnosis of Type 1 and Type 2 diabetes, current use of prescribed antidiabetic medications for at least six months, and access to AI-powered health applications. Although both Type 1 and Type 2 diabetes patients were eligible for inclusion, the final sample comprised a disproportionately higher number of individuals classified as having Type 1 diabetes. This distribution may reflect the clinical composition of the selected hospital settings, where insulin-dependent cases are more frequently managed, as well as potential variations in diagnostic classification practices across facilities. At the first stage, hospitals were stratified by location and type (public/private). At the second stage, facilities were randomly selected within each stratum. At the final stage, eligible patients were purposively recruited based on inclusion criteria.

Perceived AI-powered virtual support was assessed among participants who reported prior exposure to AI-based or mobile health support systems (e.g., reminder applications, conversational agents, or digital health platforms). Although both Type 1 and Type 2 diabetes patients were eligible for inclusion, the final sample comprised a disproportionately higher number of individuals classified as having Type 1 diabetes. This distribution may reflect the clinical composition of the selected hospital settings, where insulin-dependent cases are more frequently managed, as well as potential variations in diagnostic classification practices across facilities. It is also possible that some cases classified as Type 1 diabetes represent insulin-treated Type 2 diabetes or other forms of diabetes requiring intensive management, particularly in resource-constrained settings where advanced diagnostic differentiation may be limited. It is important to note that this measure captures participants’ subjective perceptions of AI-enabled support rather than objective usage or exposure to specific AI systems. This ensured that responses reflected experiential perceptions rather than hypothetical assumptions. Table 1 shows the socio-demographic and clinical characteristics of the participants for the main study;

**Table 1:**
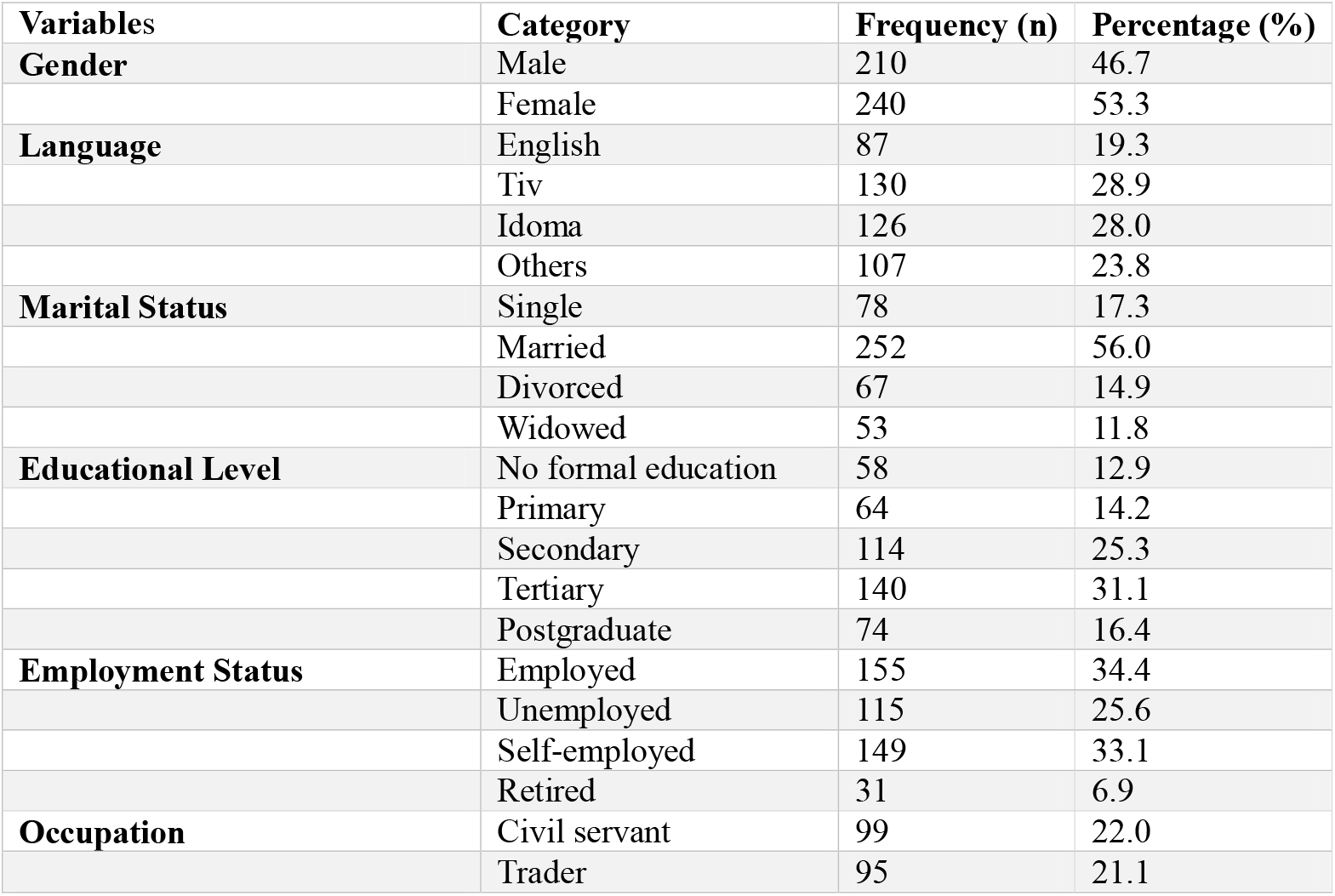
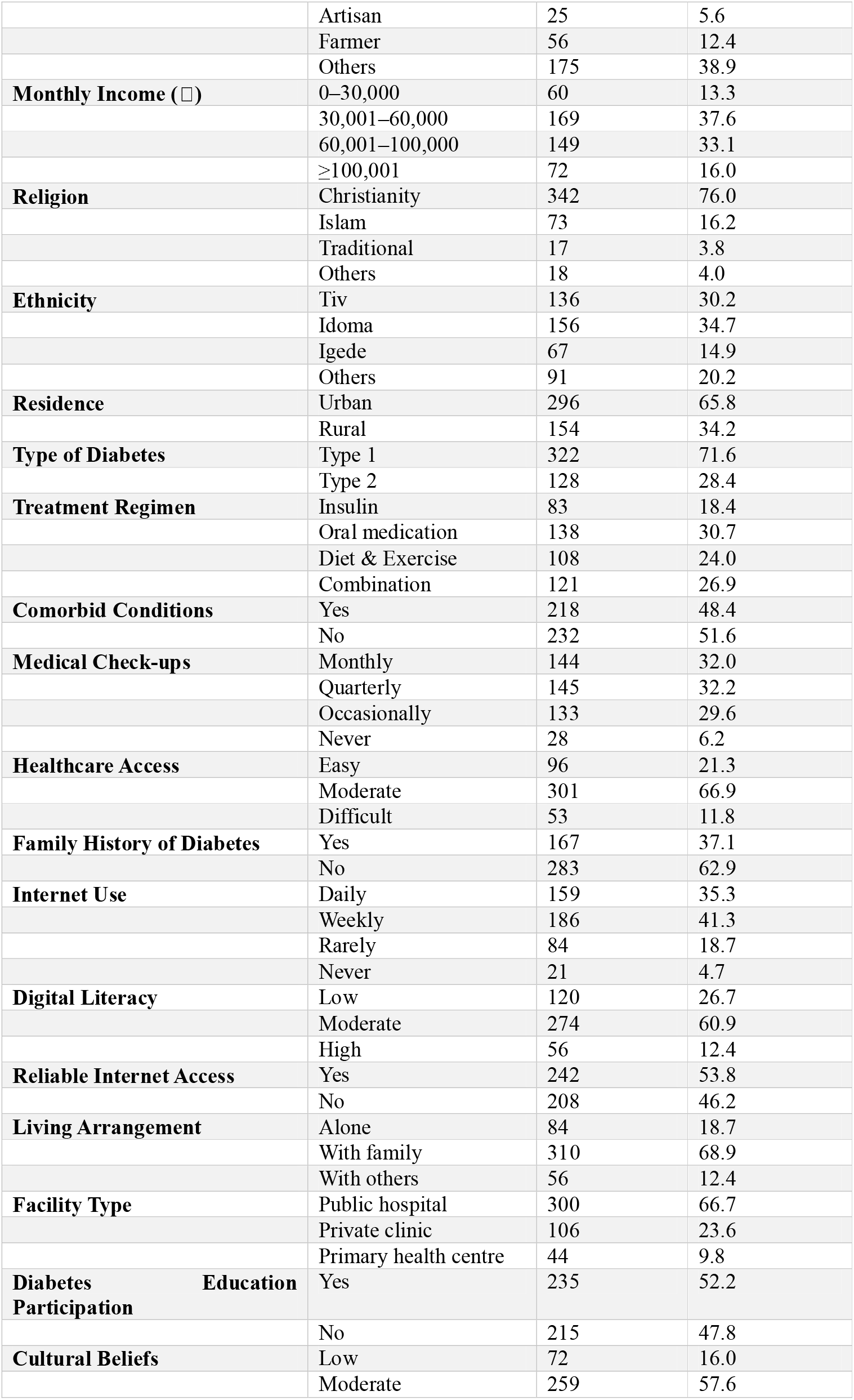
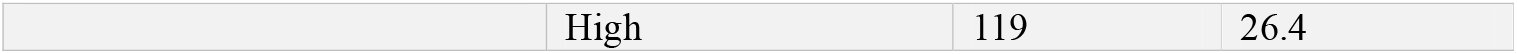
Socio-Demographic and Clinical Characteristics of Participants (N = 450)

| Variables | Category | Frequency (n) | Percentage (%) |
| --- | --- | --- | --- |
| **Gender** | Male | 210 | 46.7 |
|  | Female | 240 | 53.3 |
| **Language** | English | 87 | 19.3 |
|  | Tiv | 130 | 28.9 |
|  | Idoma | 126 | 28.0 |
|  | Others | 107 | 23.8 |
| **Marital Status** | Single | 78 | 17.3 |
|  | Married | 252 | 56.0 |
|  | Divorced | 67 | 14.9 |
|  | Widowed | 53 | 11.8 |
| **Educational Level** | No formal education | 58 | 12.9 |
|  | Primary | 64 | 14.2 |
|  | Secondary | 114 | 25.3 |
|  | Tertiary | 140 | 31.1 |
|  | Postgraduate | 74 | 16.4 |
| **Employment Status** | Employed | 155 | 34.4 |
|  | Unemployed | 115 | 25.6 |
|  | Self-employed | 149 | 33.1 |
|  | Retired | 31 | 6.9 |
| **Occupation** | Civil servant | 99 | 22.0 |
|  | Trader | 95 | 21.1 |
|  | Artisan | 25 | 5.6 |
|  | Farmer | 56 | 12.4 |
|  | Others | 175 | 38.9 |
| **Monthly Income (₦)** | 0–30,000 | 60 | 13.3 |
|  | 30,001–60,000 | 169 | 37.6 |
|  | 60,001–100,000 | 149 | 33.1 |
|  | ≥100,001 | 72 | 16.0 |
| **Religion** | Christianity | 342 | 76.0 |
|  | Islam | 73 | 16.2 |
|  | Traditional | 17 | 3.8 |
|  | Others | 18 | 4.0 |
| **Ethnicity** | Tiv | 136 | 30.2 |
|  | Idoma | 156 | 34.7 |
|  | Igede | 67 | 14.9 |
|  | Others | 91 | 20.2 |
| **Residence** | Urban | 296 | 65.8 |
|  | Rural | 154 | 34.2 |
| **Type of Diabetes** | Type 1 | 322 | 71.6 |
|  | Type 2 | 128 | 28.4 |
| **Treatment Regimen** | Insulin | 83 | 18.4 |
|  | Oral medication | 138 | 30.7 |
|  | Diet & Exercise | 108 | 24.0 |
|  | Combination | 121 | 26.9 |
| **Comorbid Conditions** | Yes | 218 | 48.4 |
|  | No | 232 | 51.6 |
| **Medical Check-ups** | Monthly | 144 | 32.0 |
|  | Quarterly | 145 | 32.2 |
|  | Occasionally | 133 | 29.6 |
|  | Never | 28 | 6.2 |
| **Healthcare Access** | Easy | 96 | 21.3 |
|  | Moderate | 301 | 66.9 |
|  | Difficult | 53 | 11.8 |
| **Family History of Diabetes** | Yes | 167 | 37.1 |
|  | No | 283 | 62.9 |
| **Internet Use** | Daily | 159 | 35.3 |
|  | Weekly | 186 | 41.3 |
|  | Rarely | 84 | 18.7 |
|  | Never | 21 | 4.7 |
| **Digital Literacy** | Low | 120 | 26.7 |
|  | Moderate | 274 | 60.9 |
|  | High | 56 | 12.4 |
| **Reliable Internet Access** | Yes | 242 | 53.8 |
|  | No | 208 | 46.2 |
| **Living Arrangement** | Alone | 84 | 18.7 |
|  | With family | 310 | 68.9 |
|  | With others | 56 | 12.4 |
| **Facility Type** | Public hospital | 300 | 66.7 |
|  | Private clinic | 106 | 23.6 |
|  | Primary health centre | 44 | 9.8 |
| **Diabetes Education Participation** | Yes | 235 | 52.2 |
|  | No | 215 | 47.8 |
| **Cultural Beliefs** | Low | 72 | 16.0 |
|  | Moderate | 259 | 57.6 |
|  | High | 119 | 26.4 |

A total of 450 participants were included in the study. The sample comprised slightly more females (53.3%) than males (46.7%). The predominant languages spoken were Tiv (28.9%) and Idoma (28.0%), followed by English (19.3%) and other languages (23.8%).

In terms of marital status, the majority of respondents were married (56.0%), while 17.3% were single, 14.9% divorced, and 11.8% widowed. Educational attainment varied, with most participants having tertiary education (31.1%), followed by secondary education (25.3%) and postgraduate qualifications (16.4%), while a smaller proportion had no formal education (12.9%) or only primary education (14.2%).

Regarding employment, 34.4% were employed, 33.1% self-employed, and 25.6% unemployed, with a minority being retired (6.9%). Occupationally, participants were largely distributed across civil service (22.0%), trading (21.1%), and other occupations (38.9%). Income distribution indicated that most respondents earned between □30,001 and □60,000 (37.6%). The majority of participants identified as Christians (76.0%), and ethnically, most were Idoma (34.7%) and Tiv (30.2%). A larger proportion resided in urban areas (65.8%) compared to rural settings (34.2%).

Clinically, most participants had Type 1 diabetes (71.6%), with common treatment approaches including oral medication (30.7%) and combination therapy (26.9%). This distribution should be interpreted with caution, as it may not reflect the population-level epidemiology of diabetes in Nigeria, where Type 2 diabetes is typically more prevalent. The observed pattern likely reflects sampling characteristics and facility-based recruitment rather than true population proportions. Approximately half of the respondents reported comorbid conditions (48.4%). Medical check-ups were commonly conducted quarterly (32.2%) or monthly (32.0%).

Access to healthcare services was largely rated as moderate (66.9%), while 37.1% reported a family history of diabetes. In terms of digital engagement, most participants used the internet weekly (41.3%) or daily (35.3%), with the majority reporting moderate digital literacy (60.9%). Slightly more than half (53.8%) had access to reliable internet services.

Most participants lived with family members (68.9%) and received care primarily from public hospitals (66.7%). Participation in diabetes education programs was reported by 52.2% of respondents. Finally, cultural beliefs about illness were predominantly moderate (57.6%), with fewer participants reporting high (26.4%) or low (16.0%) levels.

### Measures

#### Diabetics Self-Care Questionnaire (DSQ)

The DSQ was developed at the Research Institute of the Diabetes Academy Mergentheim by Schmitt (2013). It was designed to assess behaviours associated with metabolic control within common treatment regimens for type 1 and type 2, diabetes in adult patient. The rating scale was designed as a four-point Likert scale (in order to avoid a neutral response option and force a specific response) with the response options ‘applies to me very much’ (three points), ‘applies to me to a considerable degree’ (two points), ‘applies to me to some degree’ (one point), and ‘does not apply to me’ (zero points). The questionnaire allows the summation to a ‘Sum Scale’ score as well as estimation of four subscale scores. In view of their contents, the subscales were labelled Glucose Management, Dietary Control, Physical Activity, and Health-Care Use. The reliability of the SDSCA’s sum scale as all determined by Cronbach’s α coefficient to be 0.73. For the individual subscales Cronbach’s α coefficient were 0.77 for ‘Glucose Management’, 0.77 for ‘Dietary Control,’ 0.76 for ‘Physical Activities’ and 0.96 for ‘Healthcare Use’. Thus, higher scores indicate better diabetes self-management behaviour. The absence of a neutral option (no “undecided”) encourages more decisive responses and clearer interpretation.

#### Perceived AI Assistance Scale (PAISS)

This scale uses the AI assistant acceptance model examined by Zhang et al. (2021). First, they verified the positive effect of trust on the acceptance of AI assistants. Secondly, they explored the relationship with trust at the functionality and social emotional levels. Finally, they explored the mediating role of trust between functionality and social emotion and acceptance. Participants were asked to indicate their level of agreement with the following statements on the extent of using artificial intelligence from a 5-point Likert scale, with - 1 = Strongly disagree, 2 = Disagree, 3 = Uncertain, 4 = Agree, 5 = Strongly agree. Based on the technology acceptance model, for the individual subscales, Cronbach’s α coefficient of (CR = 0.906, AVE = 0.708) was found for perceived usefulness, Cronbach’s α coefficient of (CR = 0.809, AVE = 0.523) was found for perceived of ease of use, (CR = 0.926, AVE = 0.759) was found for perceived humanity, (CR = 0.743, AVE = 0.595) for perceived social interactivity, and (CR = 0.900, AVE = 0.751) for perceived social presence. However, higher scores on each subscale imply stronger perceived benefit in that domain (e.g., usefulness, ease, social connection). The presence of the ‘uncertain’ option (neutral) allows for more nuanced participant feedback.

#### Morisky Medication Adherence Scale (MMAS-8)

Medication adherence was assessed using the 8-item Morisky Medication Adherence Scale (MMAS-8), developed by Morisky et al. (2008). The MMAS-8 is a widely used and validated self-report instrument designed to measure medication-taking behaviour across chronic conditions, including diabetes. The scale consists of eight items assessing common patterns of non-adherent behaviour such as forgetfulness, carelessness, and discontinuation of medication when symptoms improve or worsen. Items 1–7 are dichotomous (Yes/No), while item 8 is rated on a 5-point Likert scale. Responses are scored and summed to produce a total adherence score ranging from 0 to 8, with higher scores indicating better medication adherence. Scores are commonly categorised as high adherence (8), medium adherence (6 to <8), and low adherence (<6), although the scale can also be treated as a continuous measure. The MMAS-8 has demonstrated good reliability and validity across diverse populations, including patients with diabetes, with reported Cronbach’s alpha coefficients ranging from 0.61 to 0.83. Its brevity, ease of administration, and strong psychometric properties make it suitable for use in clinical and research settings, particularly in low-resource environments. Permission to use the MMAS-8 was obtained from the scale developer.

### Pilot Study

The pilot involved a sample of 35 diabetic patients (both inpatients and outpatients) recruited from Benue State University Teaching Hospital (BSUTH), Makurdi, and Salem Hospital, Otukpo. This preliminary exercise also ensured that participants in the main study would not be previously exposed to the questionnaire items. Find in table 2-4 results from the pilot study.

**Table 2:** Socio-Demographic Characteristics of Pilot Study Participants (N = 35)

| Variables | Category | Frequency (n) | Percentage (%) |
| --- | --- | --- | --- |
| **Gender** | Male | 24 | 68.6 |
|  | Female | 11 | 31.4 |
| **Age (years)** | 18–24 | 3 | 8.6 |
|  | 25–34 | 5 | 14.3 |
|  | 35–44 | 8 | 22.9 |
|  | 45–54 | 9 | 25.7 |
|  | 55–60 | 10 | 28.6 |
| **Ethnicity** | Tiv | 8 | 22.9 |
|  | Idoma | 7 | 20.0 |
|  | Igede | 11 | 31.4 |
|  | Others | 9 | 25.7 |
| **Religion** | Christianity | 22 | 62.9 |
|  | Islam | 6 | 17.1 |
|  | Others | 7 | 20.0 |
| **Marital Status** | Single | 6 | 17.1 |
|  | Married | 22 | 62.9 |
|  | Divorced | 4 | 11.4 |
|  | Separated | 3 | 8.6 |
| **Educational Level** | Primary | 5 | 14.3 |
|  | Secondary (Post-primary) | 13 | 37.1 |
|  | Tertiary | 17 | 48.6 |
| **Hospital Type** | Public | 24 | 68.6 |
|  | Private | 11 | 31.4 |

**Table 3:**
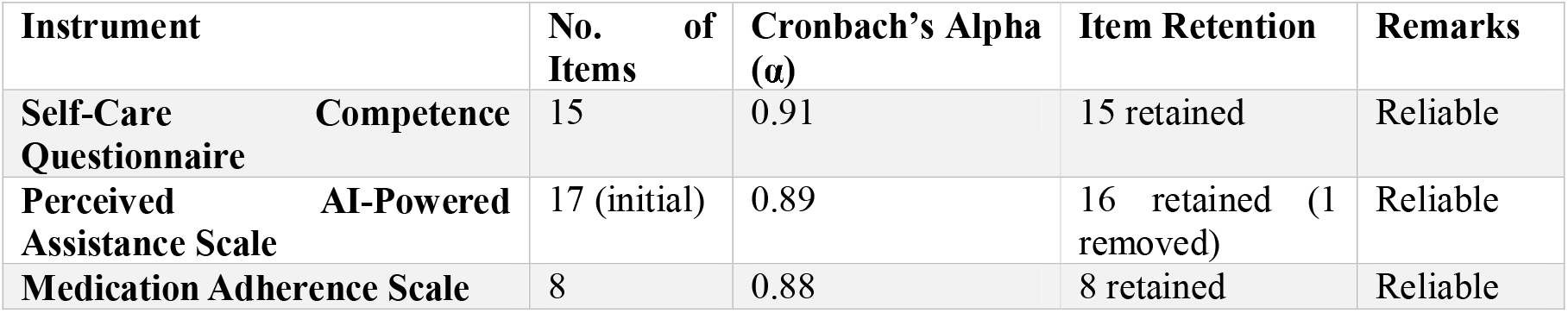
Reliability Analysis of Study Instruments.

**Table 4:** Subscale Reliability Coefficients.

| **Instruments** | **Subscale** | **Cronbach's Alpha ($\alpha$)** |
| --- | --- | --- |
| **Self-Care Competence** | Glucose Management | 0.94 |
|  | Dietary Control | 0.83 |
|  | Physical Activity | 0.76 |
|  | Healthcare Use | 0.95 |
| **AI-Powered Assistance** | Perceived Usefulness | 0.85 |
|  | Perceived Ease of Use | 0.96 |
|  | Perceived Humanity | 0.82 |
|  | Perceived Interactivity | 0.72 |
|  | Perceived Social Presence | 0.78 |

### Procedure

Prior to the commencement of fieldwork, a formal letter of introduction was obtained from the Department of Psychology, Rev. Fr. Moses Orshio Adasu University, Makurdi. This letter was presented to the management of the selected hospitals and clinics to seek permission to conduct the study. Ethical approval was subsequently obtained from the Benue State Ministry of Health and Human Services, in addition to approvals from the Research Ethics Committees and administrative authorities of the participating health facilities. These procedures ensured full compliance with established ethical standards for research involving human participants.

To facilitate efficient data collection, healthcare providers within the selected facilities were engaged as research assistants due to their familiarity with patients and local languages. The research team comprised the principal researcher and one assistant in Makurdi, alongside four assistants, supported by aides, in Otukpo and Gboko Local Government Areas. All assistants received comprehensive training on the study objectives, ethical requirements, and standardised procedures for questionnaire administration to ensure uniformity and data quality.

At each facility, the research team presented the required documentation to the Medical Director or Head of the institution and conducted brief meetings to explain the study’s purpose and procedures. Following approval, designated staff assisted in identifying and introducing eligible patients. Participants were approached in wards or clinics and provided with detailed information about the study, including assurances of voluntary participation, confidentiality, and the absence of any impact on their medical care. Written informed consent was obtained from participants, while verbal consent was documented in the presence of a witness for those unable to provide written consent.

A total of 500 questionnaires were distributed across the selected facilities. The instruments, administered in English, were completed with clarifications provided by trained assistants where necessary, while maintaining neutrality. Data collection was conducted in quiet and private settings to ensure participant comfort and confidentiality. Completed questionnaires were reviewed for completeness, and follow-up visits were conducted where required. Of the 500 questionnaires distributed, 450 were properly completed and returned, yielding a response rate of 90%. Upon completion of the data collection process, appreciation was extended to the hospital management, staff, and participants for their cooperation.

### Data Analysis

Data were analysed using both descriptive and inferential statistical techniques. Descriptive statistics, including means, standard deviations, frequencies, and percentages, were used to summarise participants’ demographic characteristics. An inter-correlation matrix was employed to examine the relationships among self-care competence, perceived AI-powered support, and medication adherence among patients with diabetes. Multiple linear regression analysis was used to test Hypotheses One and Two, while hierarchical regression analysis was employed to test Hypothesis Three.

To ensure robustness of the regression estimates, multicollinearity diagnostics were assessed using Variance Inflation Factor (VIF) and tolerance values. All predictors fell within acceptable thresholds (VIF < 5; tolerance > 0.20), indicating that multicollinearity did not bias the regression coefficients (Podsakoff et al., 2003; Kock, 2015).

## Results

Table 5 present the means, standard deviations, and pearson product–moment correlations among the study variables. Participants had a mean age of 38.28 years (SD = 13.64), with an average duration of diagnosis of 4.15 years (SD = 5.06). Medication adherence showed a moderate mean level (M = 25.92, SD = 5.58). Age was significantly and negatively correlated with key self-management behaviours, including glucose management, dietary control, and physical activities (rs ranging from −.149 to −.291, p < .01), indicating that increasing age was associated with lower engagement in these behaviours. Age was, however, positively associated with perceptions of technology-related constructs such as perceived usefulness, ease of use, humanity, and social interactivity (r ranging from .140 to .207, p < .01), as well as medication adherence (r = .099, p < .05).

**Table 5:** Inter-correlation of study variables.

| SN | Variables | Mean | Std | 1 | 2 | 3 | 4 | 5 | 6 | 7 | 8 | 9 | 10 | 11 | 12 | 13 |
| --- | --- | --- | --- | --- | --- | --- | --- | --- | --- | --- | --- | --- | --- | --- | --- | --- |
| 1 | Age | 38.28 | 13.64 | -- |  |  |  |  |  |  |  |  |  |  |  |  |
| 2 | Gender | - | - | -.314** | -- |  |  |  |  |  |  |  |  |  |  |  |
| 3 | Duration of diagnosis | 4.15 | 5.06 | .187** | -.138** | -- |  |  |  |  |  |  |  |  |  |  |
| 4 | Glucose management | 9.36 | 3.17 | -.149** | .250** | -.060 | -- |  |  |  |  |  |  |  |  |  |
| 5 | Dietary control | 7.38 | 2.34 | -.291** | .324** | -.095* | .638** | -- |  |  |  |  |  |  |  |  |
| 6 | Physical activities | 5.34 | 1.80 | -.247** | .322** | -.036 | .617** | .698** | -- |  |  |  |  |  |  |  |
| 7 | Healthcare use | 5.16 | 2.64 | -.086 | .132** | -.012 | .396** | .468** | .282** | -- |  |  |  |  |  |  |
| 8 | Perceived usefulness | 10.86 | 3.00 | .158** | -.310** | -.048 | -.461** | -.450** | -.390** | -.218** | -- |  |  |  |  |  |
| 9 | Perceived ease use | 10.65 | 3.25 | .172** | -.401** | -.039 | -.415** | -.434** | -.343** | -.235** | .708** | -- |  |  |  |  |
| 10 | Perceived humanity | 10.85 | 3.52 | .207** | -.420** | .073 | -.376** | -.368** | -.373** | -.277** | .423** | .532** | -- |  |  |  |
| 11 | Perceived social interactivity | 5.36 | 1.75 | .140** | -.300** | -.039 | -.205** | -.198** | -.227** | -.126** | .232** | .384** | .587** | -- |  |  |
| 12 | Perceived social presence | 8.02 | 2.24 | .074 | -.195** | .016 | -.266** | -.198** | -.245** | -.213** | .260** | .351** | .519** | .611** | -- |  |
| 13 | Medication adherence | 25.92 | 5.58 | .099* | -.174** | .003 | -.373** | -.276** | -.313** | -.239** | .319** | .370** | .435** | .447** | .506** | -- |

\*\* . Correlation is significant at the 0.01 level (2-tailed).
\* . Correlation is significant at the 0.05 level (2-tailed).

Gender demonstrated significant associations with most variables. Notably, gender was positively correlated with glucose management, dietary control, and physical activities (r ranging from .250 to .324, p < .01), while showing significant negative correlations with perceived usefulness, perceived ease of use, perceived humanity, and social interaction variables (r ranging from −.195 to −.420, p < .01). Duration of diagnosis was weakly related to most variables and showed only a small negative association with dietary control (r = −.095, p < .05). Strong positive inter-correlations were observed among diabetes self-management behaviours. Glucose management was highly correlated with dietary control (r = .638, p < .01) and physical activities (r = .617, p < .01). Dietary control was also strongly associated with physical activities (r = .698, p < .01) and healthcare use (r = .468, p < .01). These findings suggest that engagement in one self-management Behaviour is closely linked to engagement in others. Technology perception variables were also strongly interrelated. Perceived usefulness demonstrated a strong positive correlation with perceived ease of use (r = .708, p < .01) and moderate correlations with perceived humanity, social interactivity, and social presence (r ranging from .232 to .423, p < .01). Similarly, perceived social interactivity and perceived social presence were strongly correlated (r = .611, p < .01), indicating conceptual coherence among social perception constructs.

Medication adherence was negatively correlated with glucose management, dietary control, physical activities, and healthcare use (r ranging from −.239 to −.373, p < .01), but positively correlated with perceived usefulness (r = .319, p < .01), perceived ease of use (r = .370, p < .01), perceived humanity (r = .435, p < .01), perceived social interactivity (r = .447, p < .01), and perceived social presence (r = .506, p < .01). These results suggest that positive perceptions of technology and social interaction are associated with higher medication adherence.

Overall, the correlation matrix indicates meaningful relationships among demographic factors, self-management behaviours, technology perceptions, and medication adherence, providing support for subsequent multivariate analyses.

Table 6 presents the results of a standard multiple regression analysis examining the independent and joint prediction of medication adherence by self-care management variables, namely glucose management, dietary control, physical activities, and healthcare use. The overall regression model was statistically significant, (F(4,444) = 21.23, p < .001), indicating that the set of self-care management variables significantly predicted medication adherence. The multiple correlation coefficient was R = .401, and the model explained 16.1% of the variance in medication adherence (R^2^ = .161).

**Table 6:** Standard multiple regression showing the independent and joint prediction of Medication adherence by self-care management (Glucose management, Dietary control, Physical activities and Healthcare use)

| Variables | R | R<sup>2</sup> | F | $\beta$ | t | p-value |
| --- | --- | --- | --- | --- | --- | --- |
| (Constant) |  |  |  |  | 37.70 | 0.000 |
| Glucose management |  |  |  | -0.269 | -4.46 | <0.001* |
| Dietary control |  |  |  | 0.057 | 0.83 | 0.406 |
| Physical activities | 0.401 | 0.161 | 21.23 | -0.155 | -2.40 | 0.017* |
| Healthcare use |  |  |  | -0.115 | -2.30 | 0.022* |

*\*=Statistically significant*

Examination of the standardised regression coefficients revealed that glucose management was a significant negative predictor of medication adherence (β = −.269, t = −4.46, p < .001). This suggests that higher levels of glucose management were associated with lower levels of medication adherence when controlling for the other self-care variables in the model. Physical activities also emerged as a significant negative predictor (β = −.155, t = −2.40, p = .017), as did healthcare use (β = −.115, t = −2.30, p = .022), indicating that increased engagement in these behaviours was associated with reduced medication adherence. In contrast, dietary control did not significantly predict medication adherence (β = .057, t = 0.83, p = .406), suggesting that dietary practices did not make a unique contribution to explaining variance in medication adherence beyond the other self-care management variables.

Overall, the findings indicate that self-care management behaviours jointly contribute to the prediction of medication adherence, although their individual effects differ in magnitude and direction. The negative associations observed for glucose management, physical activities, and healthcare use highlight the complex relationship between various self-care practices and adherence to prescribed medication regimens.

These counterintuitive negative coefficients were further examined for potential statistical artefacts. Diagnostic checks indicated no evidence of multicollinearity (all VIF values < 3.0; tolerance values > 0.30), suggesting that the observed effects are unlikely to be driven by estimation instability. The pattern is therefore consistent with a suppression effect, whereby interrelated self-care behaviours share overlapping variance, resulting in negative beta weights when entered simultaneously into the regression model (Podsakoff et al., 2003).

Table 7 shows the result of a standard multiple regression analysis examining the independent and joint prediction of medication adherence by perceived attributes of AI-powered virtual assistance, including perceived usefulness, perceived ease of use, perceived humanity, perceived social interactivity, and perceived social presence. The overall regression model was statistically significant, (F(5,443)= 43.32, p < .001). The multiple correlation coefficient was R= .573, indicating a strong relationship between the predictor set and medication adherence. Collectively, the predictors accounted for 32.8% of the variance in medication adherence (R^2^ = .328).

**Table 7:** Standard multiple regression showing the independent and joint predication of Medication adherence by Perceived AI-powered virtual assistance (Perceived usefulness, perceived ease use, perceived humanity, perceived social interactivity and perceived social presence)

| Variables | R | R<sup>2</sup> | F | $\beta$ | t | p-value |
| --- | --- | --- | --- | --- | --- | --- |
| (Constant) |  |  |  |  | 11.63 | 0.000 |
| Perceived usefulness |  |  |  | 0.108 | 1.94 | 0.052* |
| Perceived ease use |  |  |  | 0.073 | 1.22 | 0.222 |
| Perceived humanity | 0.573 | 0.328 | 43.32 | 0.106 | 1.95 | 0.052* |
| Perceived social interactivity |  |  |  | 0.142 | 2.62 | 0.009* |
| Perceived social presence |  |  |  | 0.311 | 6.10 | 0.000 |

**\*=Statistically significant**

Analysis of the standardised regression coefficients showed that perceived social presence was the strongest and most significant predictor of medication adherence (β = .311, t = 6.10, p < .001), suggesting that greater feelings of social presence associated with AI-powered virtual assistance were linked to higher medication adherence. Perceived social interactivity also emerged as a significant positive predictor (β = .142, t = 2.62, p = .009), indicating that interactive features of the virtual assistant contributed uniquely to adherence Behaviour.

Perceived usefulness (β = .108, t = 1.94, p = .052) and perceived humanity (β = .106, t = 1.95, p = .052) demonstrated marginal effects, approaching statistical significance, suggesting a trend whereby greater perceptions of usefulness and human-like qualities of the AI assistant may be associated with improved medication adherence. In contrast, perceived ease of use did not significantly predict medication adherence (β = .073, t = 1.22, p = .222), indicating that ease of interaction alone did not uniquely explain variance in adherence when other perceptual factors were considered.

Overall, the findings indicate that perceptions of AI-powered virtual assistance, particularly social presence and interactivity, play a substantial role in predicting medication adherence. These results underscore the importance of socially engaging and interactive AI features in supporting adherence behaviours, beyond purely functional or usability-related attributes.

Table 8 summarises the results of a hierarchical multiple regression analysis examining the effects of self-care management behaviours and perceived AI-powered virtual assistance on medication adherence, while controlling for demographic variables (age, gender, and duration of diagnosis). Predictors were entered in three steps to assess their incremental contribution to the explanation of variance in medication adherence.

**Table 8:**
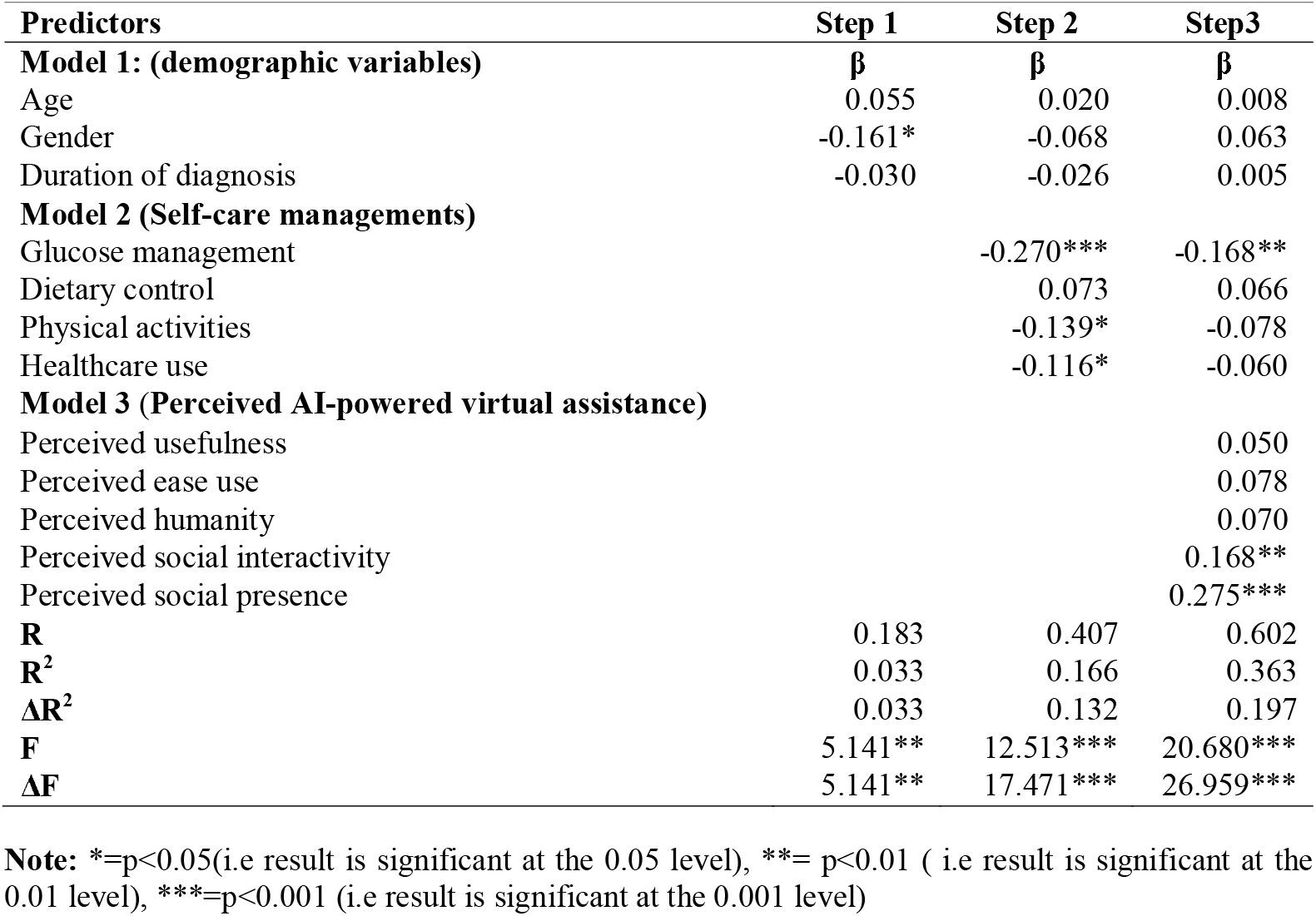
Summary of Hierarchical regression of self-care management (Glucose management, Dietary control, Physical activities and Healthcare use) and Perceived AI-powered virtual assistance (Perceived usefulness, perceived ease use, perceived humanity, perceived social interactivity and perceived social presence) on medication adherence controlling for age, gender and duration of diagnosis.

| Predictors | Step 1 | Step 2 | Step3 |
| --- | --- | --- | --- |
| **Model 1: (demographic variables)** | $\beta$ | $\beta$ | $\beta$ |
| Age | 0.055 | 0.020 | 0.008 |
| Gender | -0.161* | -0.068 | 0.063 |
| Duration of diagnosis | -0.030 | -0.026 | 0.005 |
| **Model 2 (Self-care managements)** |  |  |  |
| Glucose management |  | -0.270*** | -0.168** |
| Dietary control |  | 0.073 | 0.066 |
| Physical activities |  | -0.139* | -0.078 |
| Healthcare use |  | -0.116* | -0.060 |
| **Model 3 (Perceived AI-powered virtual assistance)** |  |  |  |
| Perceived usefulness |  |  | 0.050 |
| Perceived ease use |  |  | 0.078 |
| Perceived humanity |  |  | 0.070 |
| Perceived social interactivity |  |  | 0.168** |
| Perceived social presence |  |  | 0.275*** |
| **R** | 0.183 | 0.407 | 0.602 |
| **R<sup>2</sup>** | 0.033 | 0.166 | 0.363 |
| **$\Delta R^2$** | 0.033 | 0.132 | 0.197 |
| **F** | 5.141** | 12.513*** | 20.680*** |
| **$\Delta F$** | 5.141** | 17.471*** | 26.959*** |

**Note:** \*= $p < 0.05$ (i.e result is significant at the 0.05 level), \*\*= $p < 0.01$ ( i.e result is significant at the 0.01 level), \*\*\*= $p < 0.001$ (i.e result is significant at the 0.001 level)

In model 1, demographic variables were entered into the model. This model was statistically significant, (F = 5.141, p < .01), and explained 3.3% of the variance in medication adherence (R^2^ = .033). Among the demographic predictors, gender emerged as a significant predictor (β = −.161, p < .05), indicating a modest association with medication adherence, whereas age (β = .055) and duration of diagnosis (β = −.030) were not significant predictors at this stage.

In model 2, self-care management variables (glucose management, dietary control, physical activities, and healthcare use) were added to the model. The inclusion of these variables resulted in a significant increase in explained variance (ΔR^2^ = .132), with the model accounting for 16.6% of the total variance in medication adherence (R^2^ = .166). The change in the model was statistically significant (ΔF = 17.471, p < .001). Within this model, glucose management was a strong negative predictor of medication adherence (β = −.270, p < .001). Physical activities (β = −.139, p < .05) and healthcare use (β = −.116, p < .05) also significantly and negatively predicted medication adherence, whereas dietary control did not show a significant unique effect (β = .073). After the inclusion of self-care variables, gender was no longer a significant predictor.

In model 3, perceived AI-powered virtual assistance variables were entered into the model. This final model explained 36.3% of the variance in medication adherence (R^2^ = .363) and represented a substantial and statistically significant increase in explained variance over the previous model (ΔR^2^ = .197; ΔF = 26.959, p < .001). In this model, perceived social presence emerged as the strongest positive predictor of medication adherence (β = .275, p < .001), followed by perceived social interactivity (β = .168, p < .01). Perceived usefulness (β = .050), perceived ease of use (β = .078), and perceived humanity (β = .070) did not significantly predict medication adherence in the final model. Notably, glucose management remained a significant negative predictor (β = −.168, p < .01), although its effect size was reduced, while the effects of physical activities and healthcare use were attenuated and no longer statistically significant.

Overall, the hierarchical regression analysis demonstrates that perceived AI-powered virtual assistance contributes significantly to the prediction of medication adherence beyond demographic factors and self-care management behaviours. In particular, socially oriented AI attributes perceived social presence and social interactivity play a critical role in enhancing medication adherence, underscoring their importance in the design and implementation of AI-based health support systems.

**Diagram 3:**
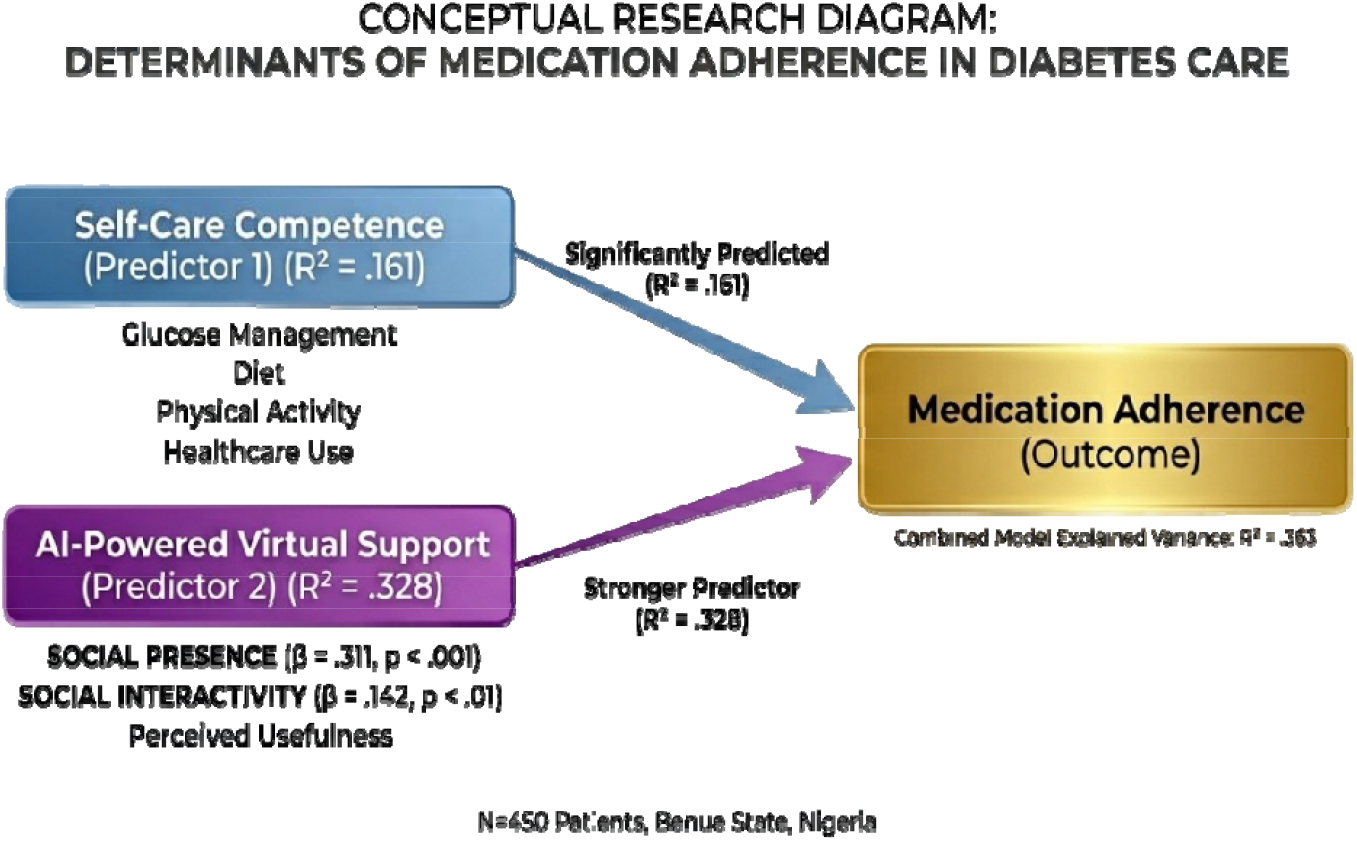
Conceptual Outcome of the Results.

### Discussion of Findings

This study examined self-care competence and perceived AI-powered virtual support as predictors of medication adherence among patients with diabetes in Benue State, Nigeria. The first hypothesis, which stated that self-care competence would significantly predict medication adherence, was supported at the joint level, although the direction of some individual predictors was unexpected. The regression results showed that self-care variables collectively explained a significant proportion of variance in adherence (R^2^ = .161). However, glucose management, physical activity, and healthcare use emerged as significant negative predictors of medication adherence, while dietary control was not significant.

The negative associations observed for glucose management, physical activity, and healthcare use require careful interpretation. Statistically, this pattern is consistent with a suppression effect, in which highly correlated self-care behaviours compete for shared variance, producing negative regression coefficients despite positive zero-order correlations. Substantively, the findings may also reflect a behavioural substitution mechanism. Patients who are highly engaged in lifestyle-based self-care (e.g., diet, exercise, glucose monitoring) may develop a heightened sense of control over their condition, which can reduce perceived reliance on pharmacological treatment.

Within the framework of the Theory of Planned Behaviour (TPB), this suggests that increased perceived behavioural control may, in some cases, weaken intention to adhere strictly to medication when alternative management strategies are prioritised. Importantly, the absence of multicollinearity concerns indicates that these effects are not merely statistical anomalies but may reflect meaningful behavioural dynamics in diabetes self-management. This finding appears counterintuitive, as self-care competence is typically associated with improved adherence. A plausible explanation is that patients who are highly engaged in non-pharmacological self-care behaviours may develop a sense of confidence or perceived control over their condition, which could inadvertently reduce strict reliance on medication (Podsakoff et al., 2003; Kock, 2015).

The findings provide a theoretical extension to TPB by showing that perceived AI-powered support may function as a form of digital subjective norm. This indicates that technologically mediated interactions can exert normative influence on health behaviour, similar to traditional social referents. Within the TPB framework, this reflects a complex interaction between perceived behavioural control and behavioural intention. When patients perceive that they can manage their condition through lifestyle practices alone, their intention to adhere strictly to medication may weaken. This finding aligns partially with Nigerian studies that highlight inconsistencies in diabetes self-management behaviours. For instance, Abdullahi et al. (2025) reported that while some patients demonstrate strong engagement in lifestyle-related self-care, this does not always translate into optimal medication adherence. Similarly, Enikuomehin et al. (2021) found uneven adherence across different domains of self-care, suggesting that competence in one domain does not necessarily predict compliance in another. The present study extends this literature by demonstrating that certain self-care behaviours may even inversely relate to medication adherence when examined simultaneously.

The second hypothesis, which proposed that perceived AI-powered virtual support would significantly predict medication adherence, was strongly supported. The observed dominance of AI-related variables over traditional self-care factors aligns with emerging global evidence suggesting that digitally mediated support systems may outperform conventional behavioural interventions in promoting adherence. This may be due to the continuous, personalised, and interactive nature of AI systems, which address real-time behavioural lapses more effectively than static self-care knowledge.

The findings revealed that Perceived AI-related variables accounted for a substantial proportion of variance, with perceived social presence and perceived social interactivity emerging as the most significant predictors. These results underscore the importance of relational and interactive features of AI systems rather than purely functional attributes such as ease of use. From the perspective of the TPB, perceived AI support can be understood as shaping both subjective norms and perceived behavioural control (Onah et al., 2024). AI systems are conceptualised as systems that simulate human-like interaction and provide continuous engagement may create a sense of social accountability and support, thereby strengthening patients’ intentions to adhere to treatment.

This is consistent with findings from Ihekoronye and Osemene (2023), who reported that mobile health interventions improve adherence by providing reminders, feedback, and a sense of connection to care. Furthermore, the strong role of social presence suggests that patients respond positively to AI systems that mimic human support, reinforcing adherence behaviours through perceived companionship and guidance. Interestingly, perceived usefulness and perceived humanity showed only marginal significance, while ease of use was not significant. This suggests that beyond basic functionality, it is the quality of interaction and emotional engagement that drives adherence. This finding contributes to the growing body of literature emphasising the social dimensions of digital health technologies in Nigeria, where interpersonal relationships and communal support systems are highly valued (Onah et al., 2024).

The third hypothesis, which stated that self-care competence and perceived AI support would jointly predict medication adherence, was also supported. The hierarchical regression analysis demonstrated that the inclusion of AI variables significantly increased the explained variance to 36.3%, indicating a strong combined effect. Notably, when AI variables were introduced, the negative effects of some self-care variables were attenuated, and only glucose management remained a significant predictor. This suggests that AI-powered support may moderate or compensate for limitations in self-care competence. Within the TPB framework, this joint effect reflects the integration of individual capabilities (self-care competence) and external influences (AI support) in shaping behavioural intention and actual behaviour. While self-care competence relates primarily to perceived behavioural control, AI support is linked to both perceived control and subjective norms, thereby strengthening adherence. The findings imply that even when patients’ self-care practices are inconsistent or misaligned, AI-powered interventions may provide corrective guidance and reinforcement.

These results are consistent with Nigerian research emphasising the role of support systems in chronic disease management. Okwuosa et al. (2022) found that perceived social support significantly predicts medication adherence among diabetic patients, highlighting the importance of external influences. The present study extends this concept by demonstrating that AI-powered virtual support can function as a digital form of social support, particularly when it incorporates interactive and socially engaging features (Onah, 2024).

The findings highlight a nuanced relationship between self-care competence and medication adherence. While self-care behaviours are essential for diabetes management, they do not operate in isolation and may sometimes produce unintended effects on medication adherence. In contrast, perceived AI-powered virtual support appears to be more consistently associated with higher medication adherence, particularly when it is linked to social interaction and presence. This underscores the need for integrated interventions that combine patient education with technologically mediated support systems.

In conclusion, this study provides empirical support for the applicability of the Theory of Planned Behaviour in understanding medication adherence among diabetic patients in Nigeria. While the Theory of Planned Behaviour provides a useful explanatory framework, the findings suggest the need to extend the model to incorporate digital and technological influences as external modifiers of behavioural intention and control, particularly in the context of AI-mediated healthcare.

Self-care competence influences adherence through perceived behavioural control, while perceived AI-powered virtual support strengthens adherence by enhancing both perceived control and subjective norms. The joint influence of these factors highlights the importance of combining traditional self-management strategies with innovative digital health solutions. For healthcare practice in Benue State and similar contexts, the findings suggest that interventions should not only focus on improving patients’ self-care skills but also leverage AI technologies that foster engagement, interaction, and a sense of social presence.

### Implications for Clinicians

The findings of this study have important implications for clinicians involved in diabetes management, particularly within resource-constrained settings such as Nigeria.

First, the significant but complex role of self-care competence suggests that clinicians should adopt a more nuanced approach when promoting self-management behaviours. While self-care practices are essential, the observed negative associations between certain self-care behaviours (e.g., glucose management and physical activity) and medication adherence indicate that patients may substitute lifestyle practices for pharmacological treatment. Clinicians should therefore emphasise the complementary—not substitutive—nature of medication and lifestyle management during patient education, ensuring that improved self-care competence does not inadvertently reduce adherence to prescribed regimens.

Second, the strong predictive value of perceived AI-powered virtual support highlights the potential clinical value of integrating digital health technologies into routine care (Onah & Gwar, 2025). In particular, features such as social presence and interactivity emerged as key drivers of adherence. This implies that clinicians should not only recommend AI-based tools but also guide patients toward platforms that provide interactive, engaging, and human-like support. Such systems can reinforce treatment plans, provide continuous monitoring, and offer real-time feedback, thereby extending care beyond the clinical setting.

Furthermore, the joint predictive effect of self-care competence and AI support suggests that optimal outcomes are achieved when patient capability is combined with external technological support. Clinicians may consider integrating AI-assisted interventions into hybrid care models, where feasible, as a complementary strategy. This approach can help bridge gaps caused by high patient loads and limited consultation time, while also enhancing personalised care.

Finally, clinicians should be trained in digital health literacy to effectively incorporate AI tools into practice and to address patient concerns regarding their use. Overall, these findings underscore the need for a shift toward more integrated, technology-enabled, and patient-centered approaches to improving medication adherence in diabetes care.

### Limitation of the study

This study has several limitations that should be considered when interpreting the findings. First, the cross-sectional design precludes causal inferences, thereby limiting the ability to establish temporal relationships among self-care competence, perceived AI-powered support, and medication adherence. Future studies employing longitudinal or experimental designs are recommended to better understand causality and directionality.

Second, the reliance on self-report measures introduces the possibility of response biases, including social desirability and recall bias, which may have influenced the accuracy of participants’ responses. Third, while the study examined perceived AI-powered virtual support, it did not incorporate objective measures of actual usage or engagement with AI-based tools. As such, the findings reflect subjective perceptions rather than verified behavioural interactions with digital health technologies.

Fourth, although the study focused on key psychological and technological predictors, potentially important confounding variables—such as socioeconomic status, comorbid health conditions, and health literacy—were not controlled, which may have influenced the observed relationships. Additionally, the study was conducted within a single state in Nigeria, which may limit the generalisability of the findings to other regions with different sociocultural and healthcare contexts.

From a procedural perspective, data collection posed practical challenges. The process of accessing, distributing, and retrieving questionnaires from patients—many of whom were experiencing varying degrees of discomfort—may have affected participation and response quality. Although efforts were made to mitigate this by providing brief psychosocial support during data collection, some degree of response burden cannot be ruled out.

Language barriers also presented a challenge, particularly among participants in rural settings who were more comfortable communicating in local dialects. This limitation was addressed through the use of trained research assistants fluent in relevant languages; however, the potential for subtle misinterpretation of questionnaire items remains.

Finally, although the study explored key dimensions of AI-powered support—such as perceived humanity and social interactivity—it did not assess the technical quality, functionality, or specific types of AI systems used by participants. Future research should incorporate more detailed and objective assessments of AI tool characteristics and usage patterns to provide a more comprehensive understanding of their role in medication adherence.

## Data Availability

All data produced in the present study are available upon reasonable request to the authors

